# Characterization of influenza vaccination recommendation across spatial scales in the United States

**DOI:** 10.1101/2021.01.19.21250118

**Authors:** Madeline C. Kuney, Casey M. Zipfel, Shweta Bansal

## Abstract

The US public health system is organized in 3 levels: national, state-level, and county-level. Public health messaging both within and across these scales may not always be consistent, and for transmissible public health threats where cases in one spatial location may impact other areas, this lack of consistency could create problems. Here, we collected and analyzed data on influenza vaccination recommendations across public health administration levels. We assess spatial heterogeneity at the county level, and analyze consistency in recommendations across spatial scales. We also compare information accessibility with influenza vaccine affordability and availability to identify factors that may be most related to vaccine uptake. We find that influenza vaccine recommendations are highly variable in both their priority group specificity and in their ease of access, and there is poor agreement across spatial scales. This lack of consistency results in a lack of clear relationship between vaccination information and vaccine uptake. This work highlights the need for greater consistency in specific, easily accessed public health information from trusted sources.

## Introduction

Seasonal influenza epidemics occur each year in temperate regions all around the world, causing devastating human and economic losses. In the United States the average annual economic loss is $87 billion, and the last moderately severe influenza season in 2016-2017 led to at least 29 million influenza-like illness (ILI) cases, over 500 thousand hospitalizations and more than 38 thousand deaths (CDC, 2020a; Molinari et al., 2007). Both of these human and economic costs could be significantly reduced with greater uptake of the influenza virus vaccine. In the United States, influenza vaccines are deemed safe and effective for all individuals over the age of six months, regardless of age, health condition or pregnancy status (A.E., C.B., & N.J., 2009). Although influenza vaccine efficacy is dependent on how well the vaccine matches the viral strains circulating in a particular season and can vary, it is thought to be between 40-80% effective at preventing influenza-like disease, and it has also been shown to significantly reduce an individual’s likelihood of developing severe disease and death (Arriola et al., 2017; Osterholm, Kelley, Sommer, & Belongia, 2012). Despite these benefits, as well as a very high and consistent safety profile, influenza vaccine uptake remains low in the United States across all age groups. Over the past decade, several policies have been introduced at both the federal and state levels to make it easier for people to access the influenza vaccine, regardless of geographic location or socioeconomic status, but little progress has been made (American Pharmacists Association, 2015; CDC, 2020c, 2020b). Between 2010 and 2018, the percentage of school-aged children receiving the influenza vaccine has improved slightly from 34.5% to 52.2% while improvement amongst adults and seniors has been much more modest, increasing from 30.5% to 34.9% and 66.6% to 68.1%, respectively (CDC, 2019). The specific factors that impact influenza vaccination rates and how they affect different groups remain largely unknown.

There are a multitude of proposed drivers of influenza vaccine uptake in the US. Some characteristics that are associated with increased influenza vaccination include having healthcare coverage, being older, white, having higher education or higher income, non-smoking, being physically active, or having poor physical health or chronic conditions (Linn, Guralnik, & Patel, 2010; Takayama, Wetmore, & Mokdad, 2012). Moving beyond characteristics, a vaccination recommendation from a healthcare professional, perceived susceptibility and severity of influenza infection, perceived vaccine effectiveness, awareness and attitudes towards vaccination are among some of the predictors of influenza vaccination (Wheelock, Thomson, & Sevdalis, 2013). It has been demonstrated that it is a combination of social and ecological drivers that result in influenza vaccine uptake, and thus successful interventions will need to target multiple factors at the interpersonal, intrapersonal, institutional, and community level (Kumar et al., 2012). Here, we examine 3 hypothesized variables the capture mechanisms related to influenza vaccination uptake in the United States: (1) availability, (2) affordability and (3) accessibility. Availability is perhaps the most basic of the three groups, as it measures how many influenza vaccines and facilities offering the influenza vaccine are within a particular area. The second of these groups, affordability, is meant to capture the financial burden of obtaining an influenza vaccine. Finally, accessibility is used in this study in reference to the accessibility of information pertaining to the influenza vaccine. This is primarily with regard to the influenza vaccine recommendation, but could also include influenza vaccine campaigns that are organized by public health entities. Especially today, with so much misinformation surrounding vaccines, access to information from official sources is important to communicate clear recommendations and to impart confidence in the influenza vaccine. Local health departments play a major role in influenza vaccination education and administration, thus we hypothesize that the more accessible and complete a health agency is able to make its information regarding the influenza vaccine, the more likely its constituents are to receive it (National Association of County and City Health Officials, 2020). Although some studies have looked at the determinants of influenza vaccination rate for specific priority groups or individual factors related to vaccination, none have yet attempted to simultaneously evaluate and compare multiple factors, across all three of these categories, and determine their comparative impact on the influenza vaccination rate of an entire population (Corace et al., 2016; Looijmans-van den Akker et al., 2009; Lu et al., 2012; Takayama et al., 2012). By integrating each of these three categories of factors into a single, comprehensive characterization of influenza vaccination in the United States, it will be possible to better identify where gaps in vaccine delivery lie and what interventions are necessary to address them.

An important consideration to be made when attempting to characterize a widespread behavior, such as influenza vaccination, is spatial scale. For example, influenza incidence data from multiple spatial scales can reveal patterns and details that are otherwise hidden or distorted when only one spatial scale is used (Bansal, Chowell, Simonsen, Vespignani, & Viboud, 2016; Lee et al., 2016). The organization of the public health system in the US heightens the importance of considering spatial scale, due to its hierarchical, decentralized structure with three main levels—national, state-level and county-level. Each level plays a critical role in public health, and specifically in promotion of the influenza vaccination. At the national level, the CDC relies on the Advisory Committee on Immunization Practices (ACIP) to determine its official influenza vaccine recommendation. State governments traditionally are the primary authority on issues related to public health, but they rely on local and county health departments to be the implementing actors. States may have a centralized, decentralized, or mixed public health governance model, which determines the relationship between the state health department and the county health departments (Meit, Kronstadt, & Brown, 2012). Here, we compare influenza vaccination factors across spatial scales. Because public health interventions can be developed by and directed towards specific spatial scales, a better understanding of the variation within and between spatial scales, and the processes that act at these scales, will allow for efforts to improve influenza vaccine uptake to become more targeted and ultimately more effective. And, we consider how lack of coordination within and among scales can impact public health. For example, promotion and uptake of the influenza vaccine in one county could significantly different from neighboring counties. This could pose significant health risks not only for those living in counties in which the influenza vaccination rate is lower, but also to those living in neighboring counties due to the deterioration of herd immunity. Therefore, one of the goals of this study will be to assess the degree of coordination, or lack thereof, both within and between the county and state levels in order to fill that knowledge gap.

The importance of spatial scale in addressing questions of public health importance and revealing mechanisms for intervention has been previously demonstrated for influenza incidence in the United States. We hypothesize that similarly important processes will be revealed for influenza vaccination in the U.S. by separating and then comparing data from different spatial scales. We propose that factors related to influenza vaccine affordability, availability and accessibility may impact influenza vaccination rate, though we expect significant variation in these factors, due to lack of coordination. To address this we 1) present data on influenza vaccine recommendations collected from county, state, and federal public health agencies 2) demonstrate the spatial distribution of variation this recommendation data 3) assess the level of agreement between different spatial scales, and 4) identify whether vaccine access, affordability, or availability play the strongest role in vaccine uptake. We encourage coordination across the public health governance for transmissible disease prevention, so that vaccination efforts can be more efficient and effective.

## Methods

### Selection of States

Ten states were selected to be included in this study: California, Colorado, Connecticut, Florida, Maryland, New Jersey, New York, North Carolina, Ohio and Washington. These ten states were chosen to ensure variation with regard to size, region, climate, degree of development (i.e. rural versus urban development), and public health governance model.

### Data Collection

- Availability: The factors selected as covariates to characterize influenza vaccine availability are the number of pharmacies and the number of primary care physicians in a given area. Both of these locations are common places where individuals can receive the influenza vaccine. The primary care physician data was obtained from the County Health Rankings and Roadmaps dataset (University of Wisconsin Population Health Institute, 2020) and the pharmacy data was obtained from the U.S. Census Bureau (US Census Bureau, 2017).
- Affordability: Although the vaccine itself does not vary drastically across the country, several factors can cause the price to vary significantly, depending on which state and county you live in. The covariate selected to capture this geographic variation is Medicare billing data from 2015 for influenza vaccinations, as reported by CMS. Another important group to consider for influenza vaccine affordability is those without health insurance. To capture this group, the rate of individuals with either private or public health insurance was also included as a covariate from County Health Rankings and Roadmap. Additionally, we included the proportion of the region that has less than a high school education as a measurement of the socioeconomic status (SES) of the population, as SES likely impacts the ability to afford vaccination, and other access issues.
- Accessibility: The website of the official health departments for a given region were used to collect two sets of data that can be used to characterize influenza vaccine information accessibility. More generally, this information is meant to capture the priorities and abilities of the public health department, as those with clear, complete, easily accessed recommendations likely play a larger role in vaccination promotion and uptake. For counties, we collected data from the County Health Department website, and for states, the State Health Department website, and for the nation, the CDC website. Data was collected from these websites as they appeared during the 2015 influenza season (September-December) using the Internet Archive WayBackMachine in order to maintain consistency with other covariates. For some county health departments, WayBackMachine did not have an archive available for the 2015 influenza season. In these cases we expanded the timeframe to 2015 – 2019 influenza seasons so that there would be data for each county, but we find that vaccination recommendations generally remain consistent over time (Figures S1 and S2). Two collected measures of information accessibility were ease and specificity.
  - Ease of access to the influenza vaccine recommendation was measured by counting the number of clicks needed to find the recommendation on the health department website for a given area. This variable had a possible range of 0 (recommendation was found on the website’s home page) to >3 (more than three clicks were needed to find the recommendation). It was also possible that a given health department did not have an influenza vaccine recommendation on its website. This study defined an influenza vaccine recommendation as a statement that includes a specific position and a directive, such as, “X County encourages all residents to get their influenza vaccine this influenza season…” Statements that were generally in favor of influenza vaccines, but did not include a directive, such as “Getting the influenza vaccine is the best way to prevent the flu,” were not counted as recommendations. In such instances, it was recorded that the website did include a “mention” of influenza or the influenza vaccine, despite having not provided a vaccine recommendation.
  - Specificity of the influenza vaccine recommendation was measured by counting the number of priority groups specifically mentioned in the recommendation. The specificity of the vaccination recommendation was considered important criteria, as it defines groups that are encouraged to receive the influenza vaccine, which communicates a clear message to the reader. The seven most common priority groups were used to define this metric, meaning that a given county or state health department could receive a specificity score within the range of 0 (no priority groups mentioned) to 7 (all priority groups specifically mentioned). The seven priority groups counted for are (1) all individuals over the age of 6 months (universal recommendation), (2) individuals over the age of 65 years, (3) women who are pregnant or plan to become pregnant during the influenza season, (4) children between the ages of 6 and 59 months, (5) healthcare workers, (6) individuals with preexisting chronic health conditions, and (7) individuals living in long term care facilities.
  - Ease and specificity scores were then combined by multiplying the two, in order to produce a single metric capable of measuring the overall information accessibility score of an influenza vaccine recommendation. This metric should provide a quantitative assessment of how accessible information regarding the influenza vaccine is to individuals at both at various administrative scales.

### Regression Model

We applied a negative binomial generalized linear mixed model to evaluate what drives vaccination at the county scale. We included a random effect for state. The response data was reports of influenza vaccination from BRFSS (Centers for Disease Control and Prevention, 2012). The potential predictor data described above were included to represent availability (primary care physicians/population size & number of pharmacies/population size), affordability (percent uninsured & average Medicare billing), and accessibility (information access score combining ease and specificity scores) of the influenza vaccine. BRFSS sample sizes were also included as a covariate to account for sampling effort. All data were z-normalized and evaluated for multicollinearity.

## Results

### Influenza vaccine recommendation ease and specificity are highly heterogeneous

Influenza vaccine recommendation ease of access (i.e. number of clicks to reach recommendation from home page) varied widely within each state, with the exception of Connecticut, where the influenza vaccine recommendation was not available on any of the county health department websites for the 2015-2016 influenza season (Figure 1). The “ease” scores were standardized to the CDC website accessibility score for the 2015-2016 influenza season, which was determined to be two “clicks”. Counties where the recommendation was equally accessible or more accessible than the CDC’s recommendation are shown in a shade of green, while counties where the recommendation was less accessible than on the CDC’s website are shown in blue. The majority of counties across these ten states had online influenza vaccine recommendations that were less accessible to their populations than the CDC.

**Figure 1.**
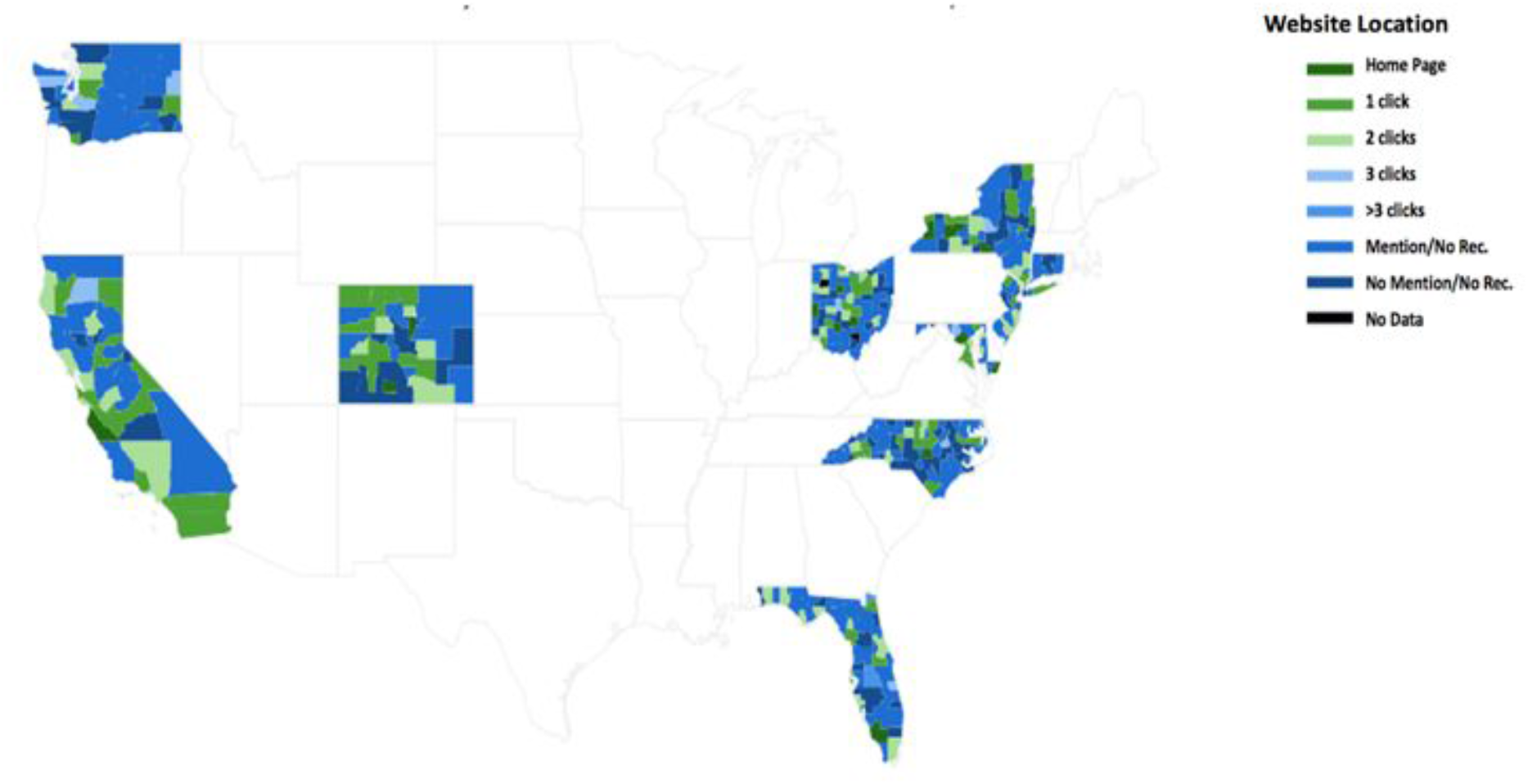
Choropleth of influenza vaccine recommendation “ease” score from health department websites by county. The color of the county indicates the number of clicks it takes from the county public health home page to reach the influenza vaccine recommendation. Mentions of influenza vaccines that were not actionable recommendations are shown as “Mention/No Rec.” Counties that did not mention influenza vaccination are shown as “No Mention/No Rec.” Counties that were not available through WayBack Machine are shown as “No Data”. Green indicates counties where the influenza vaccine recommendation was equally or more accessible on the county health department website than on the CDC website, while blue indicates counties where the recommendation was less accessible.

Counties that provided an online influenza vaccine recommendation for the 2015-2016 influenza season with a direct mention of each of the seven priority groups counted by this study are shown in green (Figure 2). All other counties are shown in a shade of blue, with darker shades indicating increasingly lower scores of recommendation specificity. Very few counties across all ten states included in this study that directly mentioned all seven priority groups in the influenza vaccine recommendation posted on their health department website for the 2015-2016 influenza season (Figure 2).

**Figure 2.**
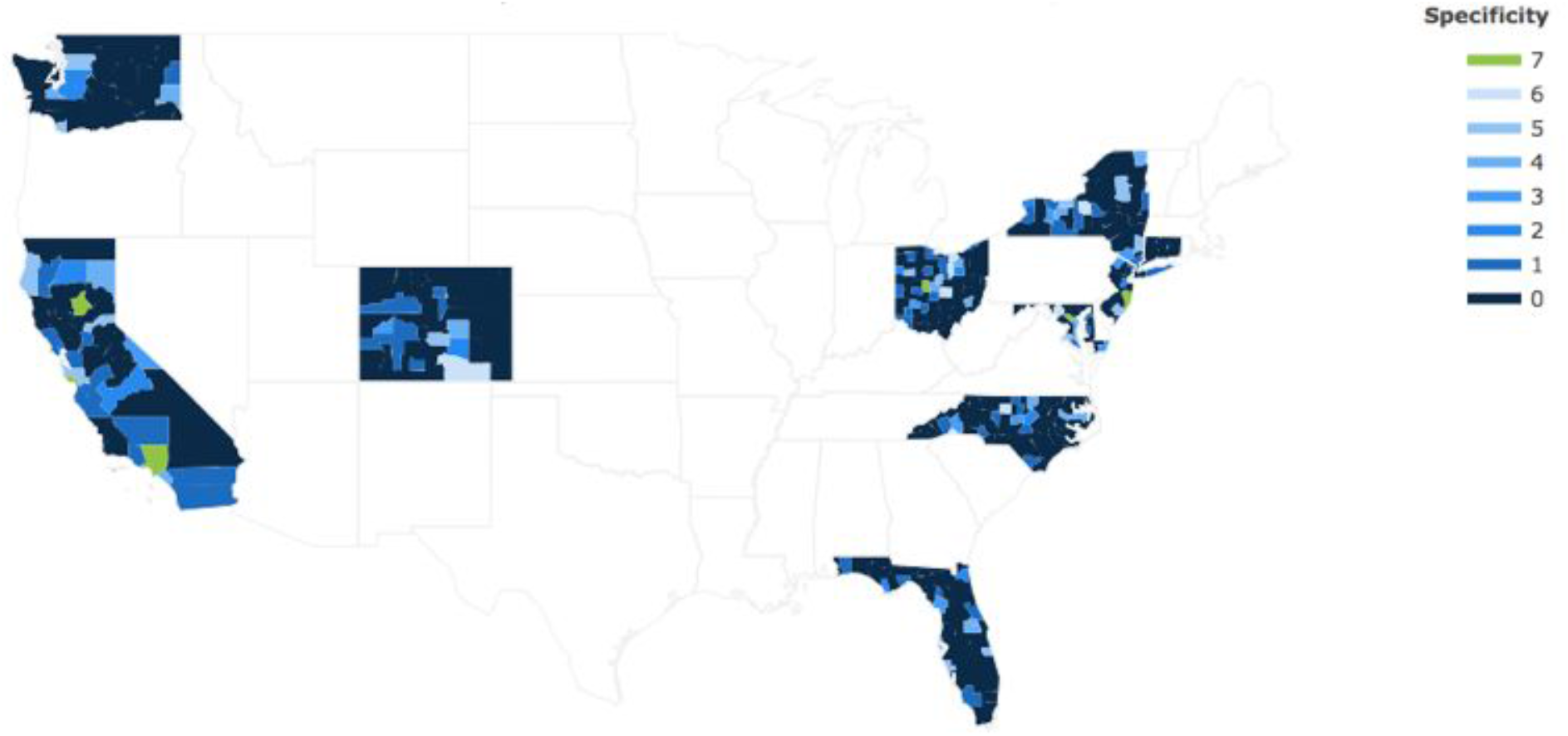
Choropleth of influenza vaccine recommendation priority group “specificity” score from health department websites by county. The color of the county indicates the number of specific priority groups included in the influenza vaccine recommendation. Green indicates counties where the influenza vaccine recommendation specifically mentioned each of the seven priority groups considered in this study, in accordance with the CDC recommendation. Blue counties have fewer priority groups mentioned.

### Influenza vaccine recommendations are spatially random and highly variable

We assessed the spatial clustering of influenza vaccine recommendation accessibility and specificity scores to evaluate if spatial proximity plays a role in recommendation consistency across counties. The clustering did not prove to be significant at the county level for most of the ten states included in this study, indicating influenza vaccine recommendation accessibility and specificity scores instead appear to be randomly distributed at the county level. North Carolina was the only state for which the degree of clustering of both recommendation ease and specificity was found to be significant. Additionally, the clustering of specificity scores was found to be significant in Ohio and the clustering of accessibility scores was significant in California (Table 1).

**Table 1.** Spatial clustering of influenza vaccine recommendation specificity and accessibility within states at the county level. Moran’s I and significance level, and the standard deviation, are shown. States for which the clustering of specificity or accessibility based on influenza vaccination rates were found to be significant are indicated in red.

| State | Specificity |  |  | Ease |  |  |
| --- | --- | --- | --- | --- | --- | --- |
|  | *Moran's I* | *p* | *Standard deviation* | *Moran's I* | *p* | *Standard deviation* |
| **California** | 0.073 | 0.128 | 1.98 | 0.156 | 0.029 | 1.95 |
| **Colorado** | 0.109 | 0.058 | 2.03 | 0.057 | 0.182 | 2.03 |
| **Connecticut** | Nan (all 0) | 0.001 | 0.52 | -0.061 | 0.408 | 0.52 |
| **Florida** | -0.04 | 0.411 | 1.25 | -0.125 | 0.069 | 1.58 |
| **Maryland** | 0.044 | 0.286 | 2.26 | 0.233 | 0.091 | 2.11 |
| **New Jersey** | 0.022 | 0.227 | 1.77 | -0.115 | 0.356 | 1.77 |
| **New York** | -0.022 | 0.47 | 2.13 | -0.027 | 0.46 | 2.13 |
| **North Carolina** | 0.1915 | 0.006 | 1.29 | 0.232 | 0.001 | 1.78 |
| **Ohio** | 0.13 | 0.023 | 1.64 | 0.089 | 0.068 | 1.90 |
| **Washington** | -0.07 | 0.374 | 1.43 | -0.119 | 0.195 | 1.54 |

The possible range of influenza vaccine recommendation accessibility scores was −1 (no data available) to 6 (recommendation found on website front page), while the possible range of recommendation specificity scores is 0 (no priority groups mentioned in recommendation) to 7 (seven priority groups mentioned). The average standard deviation for the recommendation specificity scores and accessibility scores of these ten states are 1.63 and 1.73, respectively. Only one state, Connecticut, had a standard deviation below 1.00 for both recommendation specificity and accessibility scores, indicating that there was a high degree of variability with regard to recommendation accessibility and specificity for most of the states included in this study at the county level (Table 1).

### Vaccine availability and affordability are associated with uptake

At the county-level, increased availability, characterized by higher rates of primary care physicians per person, and increased affordability, characterized by lower rates of low socio-economic status individuals, measured by education level, were associated with increased vaccination rates (Figure 3). Data coverage is also associated with increased influenza vaccination data, as expected.

**Figure 3.**
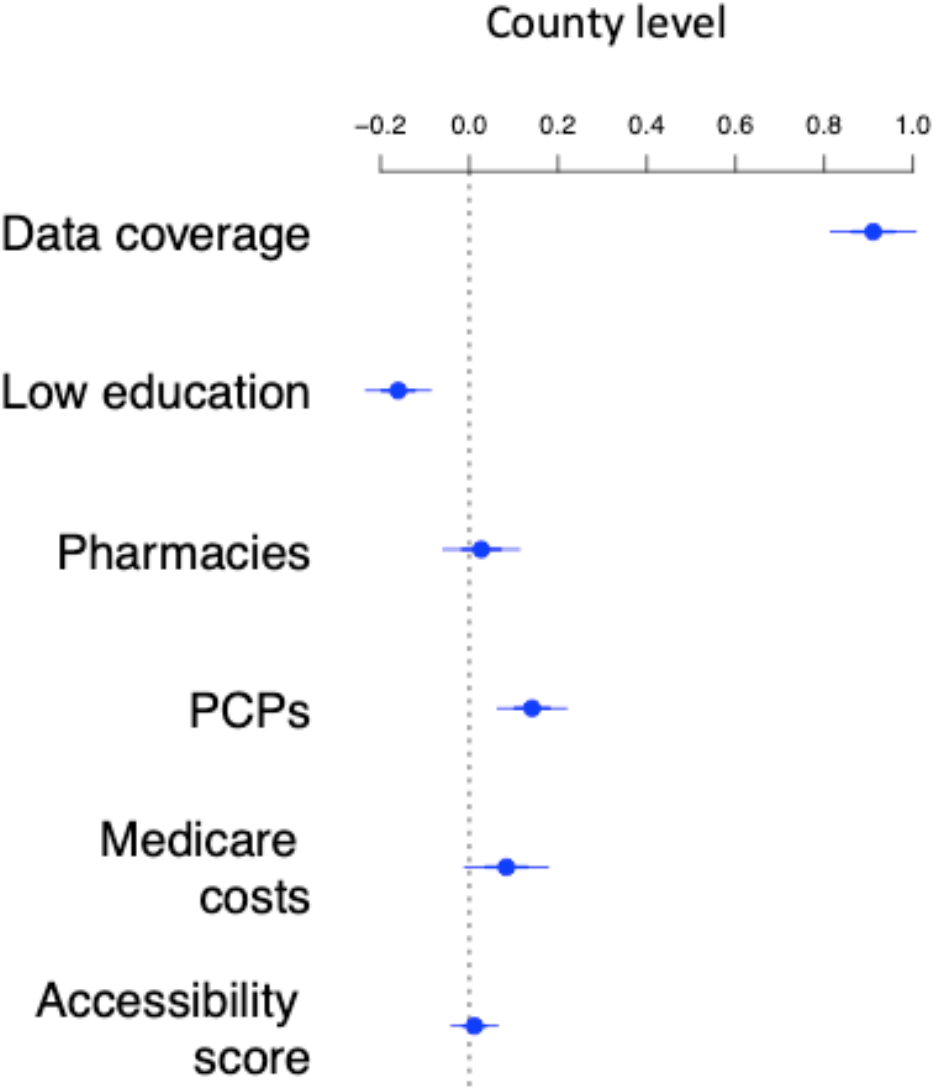
Coefficient estimates and standard errors for generalized linear models with data at the county level. Vaccine access and socioeconomic status are the primary drivers of influenza vaccination at the county level.

### There is little consistency in influenza vaccine information accessibility across scales

A comparison of county influenza vaccine recommendations with their associated state recommendation reveals that there is little consistency between county and state health departments for the ten states included in this study (Table 2). With regard to where vaccine recommendations could be found on health department websites, Maryland had the highest number of counties (5 out of 24; 20.8%) that offered recommendations that required the same number of clicks to find as the recommendation on the state health department website. Consistency between county and state health department websites with regard to vaccine specificity was slightly greater, with 49 out of 64 (76.6%) Colorado counties and 16 out of 21 (76.2%) New Jersey counties included the same number of priority groups in their official influenza vaccine recommendation as the state.

**Table 2.** Agreement between state and county official influenza vaccine recommendations. The number of counties with the same recommendation accessibility/specificity score as the official state health department recommendation were counted and divided by the total number of counties within that state to assess the degree of consistency between the state and county levels with regard to official influenza vaccine recommendations.

|  | WEBSITE LOCATION |  |  | SPECIFICITY |  |  |
| --- | --- | --- | --- | --- | --- | --- |
| State | # counties<br>in<br>agreement | Total #<br>counties | Percent<br>agreement | # counties<br>in<br>agreement | Total #<br>counties | Percent<br>agreement |
| California | 0 | 58 | 0.0% | 15 | 58 | 25.9% |
| Colorado | N/A | 64 | N/A | 49 | 64 | 76.6% |
| Connecticut | 0 | 8 | 0.0% | 0 | 8 | 0.0% |
| Florida | 4 | 67 | 6.0% | 2 | 67 | 3.0% |
| Maryland | 5 | 24 | 20.8% | 1 | 24 | 4.2% |
| New Jersey | 0 | 21 | 0.0% | 16 | 21 | 76.2% |
| New York | 11 | 62 | 17.7% | 1 | 62 | 1.6% |
| North<br>Carolina | 15 | 100 | 15% | 5 | 100 | 5.0% |
| Ohio | 3 | 88 | 3.4% | 16 | 88 | 18.2% |
| Washington | 0 | 39 | 0.0% | 0 | 39 | 0.0% |

## Discussion

The governance structure of the United States public health system results in a decentralized effort, with authority and responsibility to act distributed across multiple levels and amongst many players. While this structure is suitable for public health initiatives that are most effective when they are designed specifically for a certain group of people by local officials who are familiar with the needs and unique characteristics of that community, it can also result in inequalities and inefficiencies. This is particularly true for public health initiatives that seek to address concerns that affect the entire nation and thus require coordination and consistency at the national level, such as seasonal influenza. In order to reduce the impact of seasonal influenza and improve the influenza vaccination rate that has stagnated in the U.S. over recent years, the effect of certain factors must be understood at each of the levels from which public health is enacted.

This is the first dataset to evaluate differences in health behavior recommendations across scales. This is highly pertinent for several reasons. First, online health information plays a role in health behavior, and clear public health messaging that is easily accessible is vital (Bujnowska-Fedak & Węgierek, 2020). Several studies have shown that non-vaccinators for influenza in the US have cited a lack of understanding as a barrier to vaccination (Wheelock et al., 2013). The lack of consistency in who is specifically mentioned in vaccine recommendation (Table 2) and the lack of consistent recommendation ease of access demonstrates that this is a problem in the US. Boosting understanding and resulting vaccination through clear, consistent communication that is easily accessible from a trusted source is a worthwhile public health effort. Second, the heterogeneity in recommendation accessibility across county health departments, displayed in Figures 1 and 2, may be a marker of heterogeneity in prioritization and resource allocation of influenza vaccination. We speculate that more robust local health agencies may be reflected by more accessible public health websites. Coordinated efforts to increase influenza vaccination could span both digital and non-digital platforms; for example, those counties with complete and easily accessible recommendations are potentially also likely to promote influenza vaccination within the community with clinics and public health messaging. Further data must be collected to support this theory, but it stands to reason that strong public health messaging can be achieved both online and offline.

Variation across counties in vaccination information and uptake creates a problem for both those counties with lower vaccination rates, but also beyond. The lack of clustering seen for county-level vaccine recommendation accessibility and specificity means that these variables are likely to vary between neighboring counties and that individuals living in relatively close proximity are receiving different levels of information regarding the influenza vaccine. This is important because even if one county has a high vaccination rate due to the accessibility of the vaccine recommendation, among other possible factors, their ability to protect the most vulnerable individuals amongst them will be limited by those visiting or commuting from neighboring counties with a lower influenza vaccination rate. Furthermore, variance of recommendation accessibility between neighboring counties could lead to influenza case and fatality rate disparities, both of which would create burdens on a county health system and exacerbate pre-existing health disparities.[C1]

An intuitive way to coordinate messaging and correct the heterogeneity in recommendations is through a top down approach, where national, state, and county recommendations are consistent and easily accessed. Currently, the CDC website posts the official vaccine recommendation provided by the ACIP at the start of each influenza season, but then many county health departments either use a far less specific, or informative, version of that recommendation or post it on their website where it is much more difficult to locate (Figure 1 and 2). This lack of coordination may also be why affordability and availability of influenza vaccination appear to play a larger role in vaccine uptake at the county level. Information accessibility has been shown to be a barrier to vaccination in prior studies. We believe the lack of coordination and resulting randomness in the relationship between recommendations and vaccinations is driven by this lack of coordination. When there are different recommendations dependent on what administrative-scale is observed, a clear effect of improved information resulting in improved vaccine uptake would be obscured.

This study has also revealed that there is a high level of variability at both the county and state levels with regard to the factors that affect influenza vaccination rate. While some variability within the U.S. public health structure is assured by its decentralized governance structure, influenza vaccination is one example of a public health concern that does not necessarily benefit from that model. Because public health authority is so spread out, the accessibility, availability and affordability of the influenza vaccine may vary drastically across both the county and state levels, depending on whether or not a specific state or county health department has the resources and inclination to consider influenza vaccination a priority. This results in an overall lower national vaccination rate and limits the ability of states and counties with high vaccination rates to prevent influenza cases and fatalities. According to a study conducted by NORC in 2012, the majority of state health departments already collaborate with county health departments by “exchanging information,” working together on activities or projects,” and “providing financial resources” (Meit et al., 2012). This suggests that in many cases there are mechanisms already in place through which states can work with county health departments to improve coordination and consistency between the state and county level as well as across the county level with regard to the factors of influenza vaccination.

One benefit that does come of the U.S.’s decentralized approach to public health is that health officials can address concerns from each of the three levels of governance, depending on which level will have the greatest effect. Given this, there are four recommendations that can be made based on the findings of this study for how best to improve the U.S. influenza vaccination rate:

1. **State health departments should first focus on making their influenza vaccine recommendation more accessible on their official website and more specific, to include all priority and vulnerable groups**. Consistency can be ensured by instructing states to use the same vaccine recommendation found on the CDC’s website. This should be the first step towards improving the influenza vaccination rate from the state level because 1) it was found to have a significant positive correlation with vaccination rate and 2) it would not require any additional resources or funding from the state.
2. **County health departments should assess whether vaccine affordability is a barrier to vaccination amongst their residents**. Because affordability was determined to be a significant factor for determining vaccination rate at the county level, counties where a significant portion of the population belongs to a lower socioeconomic class may be able to effectively increase vaccination rates by instituting programs that provide free or subsidized vaccines.
3. **County health departments should assess whether vaccine availability is a barrier to vaccination**. Because increased availability (measured by number of PCPs) was found to significantly correlate with increased influenza vaccination rate, counties with a limited number of PCPs should focus their resources towards making the vaccine more available at other locations throughout the community.
4. **State and county health departments should incorporate efforts to improve influenza vaccination rate into pre-existing mechanisms of collaboration**. By using the methods that state and county health departments already use to communicate, coordination and consistency can be achieved for new initiatives to improve influenza vaccination rates, which will ultimately make these efforts more effective and decrease spatial disparities in influenza case and fatality counts.

Just as looking at influenza incidence from multiple spatial scales can reveal previously unknown patterns and insights, considering the factors that affect the influenza vaccination rate from the three primary levels of the U.S. public health system can help identify the most effective methods to improve it. Using this approach may ultimately help to reduce the human and economic loss experienced by the U.S. every influenza season.

## Data Availability

All data were gathered from publicly available sources and are referenced in the manuscript. Compiled county and state recommendation data is available at https://github.com/bansallab/fluvacc_acrossscales

## Notes

### Competing Interest Statement

The authors have declared no competing interest.

### Funding Statement

Research reported in this publication was supported by the National Institute Of General Medical Sciences of the National Institutes of Health under Award Number R01GM123007. The content is solely the responsibility of the authors and does not necessarily represent the official views of the National Institutes of Health. We also acknowledge support from the PhRMA Foundation.

