## Supplement for "Characterization of influenza vaccination recommendation across spatial scales in the United States"

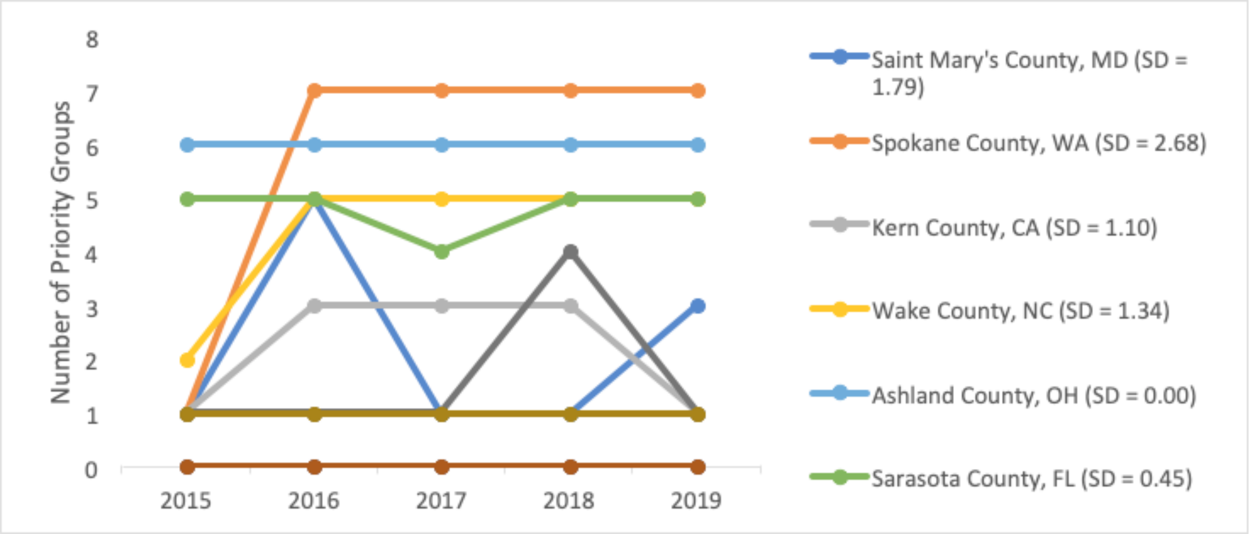


Figure S1. Temporal continuity (2015-2019) for influenza vaccination specificity at the county level.

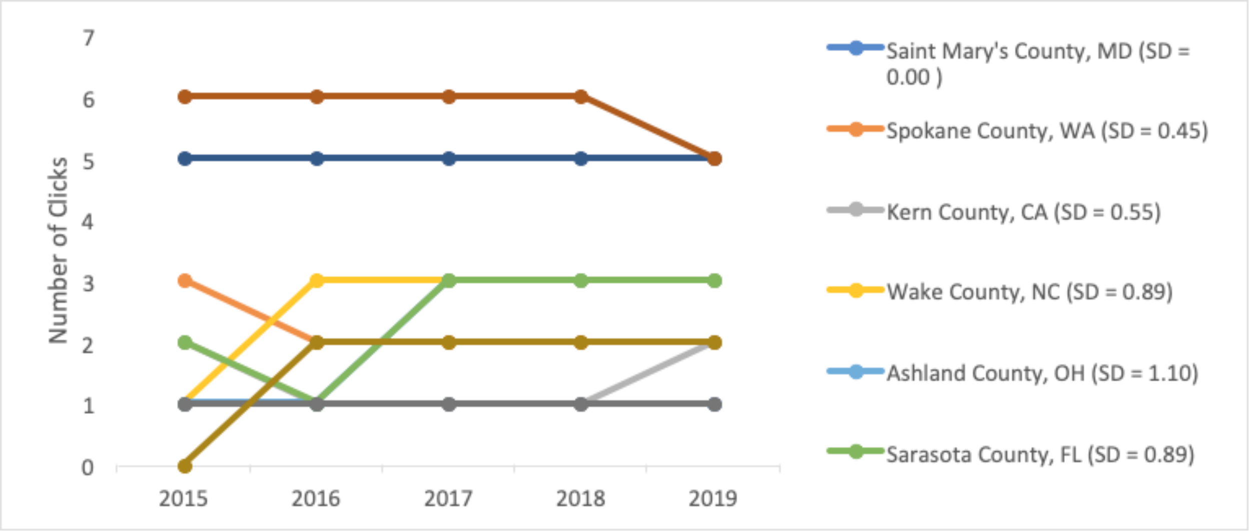


Figure S2. Temporal continuity (2015-2019) for influenza vaccination accessibility at the county level.
